# Genomic epidemiology and emerging antimicrobial resistance profiles of *Salmonella* Paratyphi A in returning travellers to Australia

**DOI:** 10.64898/2026.08.20.26360952

**Authors:** Christopher H. Connor, Ryan R. Wick, Mona L. Taouk, Jessica Barnden, Sally Dougall, Jane McAllister, Louise M. Judd, Karolina Mercoulia, Benjamin P. Howden, Danielle J. Ingle

**Affiliations:** Department of Microbiology & Immunology at the Peter Doherty Institute for Infection & Immunity, University of Melbourne, Melbourne, Victoria, Australia; Centre for Pathogen Genomics, University of Melbourne, Melbourne, Victoria, Australia; Microbiological Diagnostic Unit Public Health Laboratory, Department of Microbiology & Immunology at the Peter Doherty Institute for Infection & Immunity, University of Melbourne, Melbourne, Victoria, Australia; WHO Collaborating Centre for Foodborne Disease Surveillance and Genomics at the Microbiological Diagnostic Unit Public Health Laboratory, University of Melbourne, Melbourne, Victoria, Australia; Department of Health Victoria, Melbourne, Victoria, Australia; Department of Infectious Diseases & Immunology, Austin Health, Heidelberg, Victoria, Australia

**Keywords:** Pathogen genomics, surveillance, enteric fever, antimicrobial resistance, surface antigens

## Abstract

Enteric fever is endemic to many low- and middle-income countries (LMICs), particularly those in sub-Saharan Africa, South and South-East Asia. The causative agents are typhoidal serovars of *Salmonella enterica,* including Typhi (*S.* Typhi) and Paratyphi A (SPA). There are no vaccines currently licensed for SPA, leaving antimicrobials as the only therapeutic option. Multi-drug resistance (MDR) *S.* Typhi is increasingly prevalent, but to date has not been detected in SPA. In Australia, cases of SPA are notifiable. Here we report on the genomic epidemiology of 208 cases of SPA in returned travellers to Australia, and their close contacts, from 2018 to 2025. A total of 15 unique genotypes were detected, and these were correlated with geographical regions of reported travel. There was a low incidence of antimicrobial resistance with only a single isolate carrying acquired resistance genes. Mutations in quinolone resistance determining regions were common across the genotypes, detected in 95.7% of isolates. A single isolate in a traveller returning from India was resistant to several first line antibiotics including: ampicillin, amoxicillin plus clavulanic acid, ceftriaxone, azithromycin and ciprofloxacin. The isolate carried a plasmid encoding an extended spectrum beta-lactamase (*bla*_CTX-M-231_), two macrolide resistance genes (*mphA* and *ermB*) and a quinolone resistance gene (*qnrS1*). Elements of the pangenome were explored, with stable maintenance of small plasmids encoding hypothetical proteins detected in four genotypes. Copy number variation in genes encoding surface antigen biosynthesis genes were detected in six genotypes. These biosynthesis genes are targets for one of the two SPA vaccines in development, and the potential variation in surface antigens could have implications for vaccine efficacy. Linking epidemiological data with genomic studies of SPA provides an opportunity to improve understanding of the emergence, spread and risk of drug-resistant SPA infections, and to better inform empirical treatment guidelines in returned travellers.

**AUTHOR SUMMARY:** Enteric fever is a disease characterised by a prolonged fever, fatigue and diarrhoea. The burden of disease is disproportionately experienced in children <5years in low-and middle-income countries. One of the causative agents is *Salmonella enterica* serovar Paratyphi A (SPA). There is no licensed vaccine for SPA, leaving antibiotics as the only treatment option. Increasing antimicrobial resistance (AMR) in SPA is of concern. Enteric fever is not endemic in Australia but can be acquired by individuals travelling to high-risk regions, such as Sub-Saharan Africa, South and South-East Asia. Using genomics on SPA isolates collected from returned travellers allows for informal surveillance of disease across a range of geographical regions. Rates of AMR were low, however there was a single case with a multi-drug-resistant profile. Concerningly this resistance could be easily spread between pathogens, highlighting the need to continued surveillance of this disease. Further, differences in genes involved in surface antigens was detected, which may have implications for the development of vaccines.

## INTRODUCTION

Enteric fever is a systemic bacterial infection that presents with symptoms such as a prolonged fever, fatigue and diarrhoea. In severe cases the disease can progress to intestinal perforation, sepsis and death (1). In 2021 there were 9.3 million cases of enteric fever globally and 107.5 thousand associated deaths, children under the age of 5 are particularly vulnerable, accounting for 40% of deaths globally (2). The disease is transmitted via the faecal oral route (3). While improvements in safe drinking water, sanitation and hygiene (WASH initiatives) have resulted in the elimination of enteric fever in many countries, it is still endemic in regions such as Sub-Saharan Africa, South and South-East Asia (2).

Enteric fever is caused by *Salmonella enterica* serovars Typhi (*S*. Typhi), Paratyphi A (SPA), Paratyphi B or Paratyphi C which cause typhoid and paratyphoid fevers respectively. *S.* Typhi accounts for most cases of enteric fever, causing an estimated 76.3% of infections globally, followed by SPA (4). While typhoid and paratyphoid fever are caused by different pathogens, they are clinically indistinguishable (5) and first line treatment for both remains antibiotic therapy. As older generations of antibiotics, such as chloramphenicol, are considered to be ineffective commonly recommended antibiotics now include ciprofloxacin (quinolone), ceftriaxone (beta-lactam) and azithromycin (macrolide) (6). In Australia, the recommended therapeutic for patients requiring hospitalisation (and have not travelled to Pakistan) is a third-generation cephalosporin or azithromycin (7).

Unlike *S.* Typhi, there is currently limited AMR in SPA. In serovars of *Salmonella enterica,* resistance to quinolone antibiotics, such as ciprofloxacin and nalidixic acid, is typically mediated by triple point mutations in quinolone resistance determining regions (QRDRs) which encode genes including *gyrA*, *gyrB*, *parC* and *parE.* Alternatively, resistance can be mediated by a QRDR point mutation and an acquired resistance gene such as *qnrS* (8,9). Resistance to beta-lactam antibiotics is conferred by Extended Spectrum Beta Lactamase (ESBL) genes, such as *bla*_CTX-M-15,_ while resistance to azithromycin is due to point mutations in the efflux pump *acrB* (10–12). However, there is concern that AMR could increase in SPA over the coming years, particularly as multi-drug resistant (MDR), and extensively drug resistant (XDR) *S.* Typhi have been increasingly reported (13–17).

The emergence of MDR, and now XDR *S*. Typhi, has spurred the development and deployment of a vaccines against enteric fever (13). The World Health Organisation (WHO) has recommended three classes of vaccine to protect against enteric fever; typhoid conjugate vaccines (TCV), Ty21a (an attenuated *S*. Typhi strain) and Vi capsular polysaccharide vaccines (ViCPS) (18,19). These vaccines have been deployed and showed evidence of benefit (20), whilst early reports indicate no adaptive changes in the *S*. Typhi population (21). Unfortunately, SPA lacks the capsular Vi antigen targeted by these vaccines and as such, they provide no protection against SPA infections (5,22). There are several vaccine candidates for SPA in varying stages of development, the most advanced of these is a live attenuated vaccine in phase 2 trials (CVD1902) (23,24). The deployment of a vaccine against *S*. Typhi whilst SPA is not covered has led some to speculate that cases of SPA will increase, with some early evidence of this occurring (25,26). Surveillance of SPA is important for monitoring changes in incidence in response to vaccine deployment, emergence and spread of AMR as well as identification of surface antigens for future vaccine targets.

A genotyping scheme, Paratype, has been previously developed to provide a genomic framework and standardised nomenclature for SPA (27). This scheme enables enhanced genomic surveillance of SPA populations, facilitating identification of expanding genotypes. Together with additional genomic analyses and epidemiological data, it enables the detection of known and emerging AMR patterns, and geographical associations with genotypes. This mirrors the approach successfully implemented for *S*. Typhi where Typhi Mykrobe enables rapid and robust genotyping of *S.* Typhi direct from reads (15) and the genotyping scheme provided the basis for global meta-analyses (13). Surface antigens are important for SPA virulence and as potential vaccine targets. The synthesis and transport of the O-antigen in SPA is controlled by genes in the *rfb* locus (22,28), consequently Paratype reports mutations in *rfbS* (*prt*),*rfbB* (*rmlB*), *rfbC* (*rmlC*), *rfbD* (*rmlD*) and *rfbX* (*wzx*) (27).

SPA is not endemic in Australia with cases typically only seen in returning travellers, or their close contacts. The disease is a Nationally Notifiable Disease requiring that all laboratory confirmed cases are notified to the local state or territory Department of Health which are in turn reported to the Australian Centre for Disease Control via the National Notifiable Diseases Surveillance System (NNDSS) (29). In the Australian state of Victoria, the Microbiological Diagnostic Unit Public Health Laboratory (MDU PHL) serves as the reference laboratory for all isolates collected in the state. Since mid-2018 MDU PHL has been routinely sequencing all *Salmonella* isolates received. Here we use the genomic data from 208 isolates received between 2018 and 2025, representing an unbiased collection of SPA isolates. We sought to characterise the genomic epidemiology, key genotypes, AMR profiles, and differences in surface antigens of SPA collected in Australia. This serves as informal sentinel surveillance of regions frequented by returned travellers, as has been previously applied to *S*. Typhi (30), providing insight in the SPA circulating in Australia and the surrounding region.

## METHODS

### Ethics

Data were collected in accordance with the Victorian Public Health and Wellbeing Act 2008. Ethical approval was received from the University of Melbourne Human Research Ethics Committee (reference number 2024-30320-59894-3)

### National case notification, total travellers and metadata collection

All microbiologically confirmed cases of paratyphoid fever are nationally notifiable through Australia’s National Notifiable Disease Surveillance System (NNDSS). All isolates collected within the state of Victoria are referred to the Microbiological Diagnostic Unit Public Health Lab (MDU PHL). Notifications received by jurisdiction to the NNDSS system were collected from the data dashboard (https://nindss.health.gov.au/pbi-dashboard/) filtering disease name to ‘Paratyphoid’. The period was filtered to between July 2018 and September 2025, inclusive. The total number of returning travellers was accessed from the Australian Bureau of Statistics (ABS) (https://www.abs.gov.au/statistics/industry/tourism-and-transport/overseas-arrivals-and-departures-australia/latest-release) using the data explorer datasets. Patient travel history was collected by the Victorian Department of Health.

### Isolate purification, MIC testing and whole genome sequencing

Samples referred to MDU PHL are sub-cultured onto a Nutrient Agar (NA) plate and incubated at 37°C for 18–24 hr. For antibiotic susceptibility testing, values were determined using the broth microdilution method with the Sensititre NARMS Gram Negative CMV5AGNF Plate (Thermo Fisher Scientific). Values were determined and interpreted with EUCAST methodology (2025 breakpoints). For whole genome sequencing a single colony was harvested with a 1 μl sterile inoculating loop and emulsified into 200 μl lysis buffer, genomic DNA was then extracted using the QIAsymphony™ DSP DNA Virus/Pathogen Kit (Qiagen) according to manufacturer’s instructions. Sequencing is then performed using the Illumina Nextera XT DNA Library Preparation kit with 150 bp paired-end reads following manufacturer’s instructions.

### Short read sequencing quality control and species identification

Illumina sequencing reads were analysed in a Nextflow v25.04.6 pipeline (31). Sequence reads were trimmed with Fastp v1.0.1 (32) using a reverse sliding window of 3bp, minimum quality threshold of 20 and a minimum length of 36bp. Samples where more than 70% of their sequencing reads did not pass trimming were discarded, isolates with less than 30x coverage were also removed. Trimmed reads were used for species identification with Kraken2 v2.1.6 (33) and the Genome Taxonomy Database (GTDB) release 214 (34), only pure isolates matching to *S. enterica* were kept. Trimmed reads were assembled using Shovill 1.1.0 (35) with a minimum contig length filter of 200bp, assembly quality was assessed with Quast v5.3.0 (36). Isolates with more than 500 contigs, or a genome size that deviated by more than 20% from the SPA reference genome (AKU_12601, GCF_000026565.1) were excluded.

### Genomic typing, antimicrobial resistance gene and plasmid detection

Multi-locus sequence typing was performed with mlst v2.23.0 (37,38) using the Achtman 7 gene scheme (39). *Salmonella* specific serotypes were predicted with Sistr v1.1.3 (40) and genotypes were assigned with Paratype v1.1 (27) using the trimmed sequencing reads as input. AMR genes were detected in the assemblies with AbriTAMR v1.0.19 using the *Salmonella* species flag (41), and plasmid detection was performed with MOB-Suite v3.1.9 with default parameters (42).

### Phylogenetic analysis

Trimmed Illumina reads for each included sample were mapped to the standard SPA AKU12601 reference genome (GCF_000026565.1) with Snippy v4.6.0 (43) using a minimum coverage threshold of 10 and a minimum fraction threshold of 0.9. The SNP calls were combined into a core SNP alignment with snippy-core, while masking phage regions predicted by PHASTEST v3.0(44). Recombination was detected and removed from the core SNP alignment using Gubbins v3.4.3 (45) with the RAxML-NG tree builder (46), a GTRGAMMA model, a recombination convergence method and 1000 bootstraps. The tree was visualised in R v4.5.0 with ggtree v3.17.0 (47,48).

### SNP distance analysis

To measure the similarity or dissimilarity of specific isolates, such as repeat samples collected from the same patient, a SNP distance between the isolates was calculated. Isolates were mapped to the AKU12601 reference genome with Snippy as described above, the resulting SNP calls for the specific pair of isolates to be compared were then used to construct an isolate pair specific core genome alignment with snippy-core. The SNP distance between the two isolates was then calculated with snp-dists v0.8.2 (49) with default parameters.

### Long-read sequencing and assembly

Nine isolates were sequenced with Oxford Nanopore Technologies (ONT) long-read sequencing to obtain fully resolved plasmid and *rfb* locus sequences (**Supplementary Table 1**). Five isolates were sequenced on R10.4.1 MinION flow cells. DNA was extracted using QIAGEN DNeasy Blood and Tissue Kit according to the manufacturer’s instructions (QIAGEN) using lysozyme and proteinase K without size selection. Sequencing libraries were made with the SQK-RBK114.96 kit and basecalling for the ONT R10 sequencing was performed using Dorado with the sup@v4.3.0 basecalling model. An additional four isolates were sequenced on a GridION Mk1 (Oxford Nanopore Technologies) using R9 Flow cells (FLO-MIN106D, Oxford Nanopore Technologies) (**Supplementary Table 1**). DNA was extracted using the QIAsymphony™ DSP DNA Virus/Pathogen Kit (Qiagen) and libraries prepared with the Ligation Sequencing Kit (Product Code SQK-LSK110, Oxford Nanopore Technologies). Basecalling for the ONT R9 sequencing was performed using Dorado v7.9.8 with the hac@v3.3 basecalling model. The ONT R10 data were assembled using Autocycler v0.5.2 (50), followed by Illumina-read polishing with Polypolish v0.6.0 (51) and Pypolca v0.4.0 (52). The ONT R9 data were hybrid assembled with Illumina reads using Unicycler v0.5.1 (53).

### Plasmid characterisation and comparison with public data

Plasmid sequences in this study were compared to publicly available sequences to identify prior reports, organisms of isolation and geographic spread. The plasmid sequences generated from hybrid assembly were used as query sequences to search against the NCBI non-redundant core nucleotide database with BLASTn using default parameters (megablast) (https://blast.ncbi.nlm.nih.gov/Blast.cgi). Presence of plasmid sequences in our dataset was confirmed using the command line version of BLASTn v2.17.0 (54), again using the plasmid sequences from hybrid assemblies as query sequences with default parameters. To capture full length hits whilst avoiding hit fragmentation, query circular plasmid sequences were doubled and concatenated end to end. Hits were filtered to those with greater than 90% coverage (adjusting for doubled query length) and 90% identity.

### *Rfb* locus alignment

The nucleotide sequence of the *rfb* locus (860063 to 884690) was extracted from the SPA AKU_12601 reference genome (GCF_000026565.1). The extracted locus was used as the query sequence for a BLASTn v2.17.0 search against our complete genome assemblies (55), to identify the position of the *rfb* locus in each assembly. Individual loci were extracted from the complete genomes using SeqKit v2.9.0 and were combined into a single FASTA file (56). The loci were then aligned against each other using Minimap2 v2.30-r1287 (57), and genes were annotated using Bakta v1.11.4 with the full Bakta database v6.0 (58). The results were visualized in R v4.5.0 using the gggenomes v1.1.0 package (47,59). The 2,885bp tandem duplication was extracted from the AKU_12601 reference genome (870332 to 873216) using SeqKit. Copy number in the closed genomes was determined with BLASTn.

### Data availability

The SPA genomes have been uploaded to the MDU PHL BioProject PRJNA857539. Details for the individual isolates including accessions, ONT sequencing technology, year of collection, genotype and AMR profile are reported in **Supplementary Table 1.**

## RESULTS

### Incidence and origins of SPA in travellers

Between July 2018 and September 2025 there were 208 notifications of Paratyphoid disease to the National Notifiable Diseases Surveillance System (NNDSS) from Victoria (**Figure 1A).** During the same period the Microbiological Diagnostic Unit (MDU) at the Victorian Public Health Lab received a total of 301 bacterial isolates collected from 207 unique patients. Only the first isolate received per patient was used for analysis. The exception was for a single patient who contracted paratyphoid twice during the study period with a 6-year gap between samples, and while both cases were acquired from travel to Pakistan, there were 204 SNPs between the isolates, bringing the dataset to 208 unique cases (**Figure 1A).** There was a notable drop in cases between March 2020 and January 2022, corresponding to the closure of Australian borders due to the COVID-19 pandemic, consistent with observations made from other bacterial associated pathogens (60,61). Excluding the border closure period, there was an average of 8.87 cases per quarter. There was no change in the rate of cases pre- and post-border closures. Case incidence followed a seasonal pattern with incidence highest between January and March (mean of 15.0 cases per quarter) corresponding to the Australian summer and lowest in the winter months between July and September (mean of 4.17 cases per quarter).

**Figure 1.**
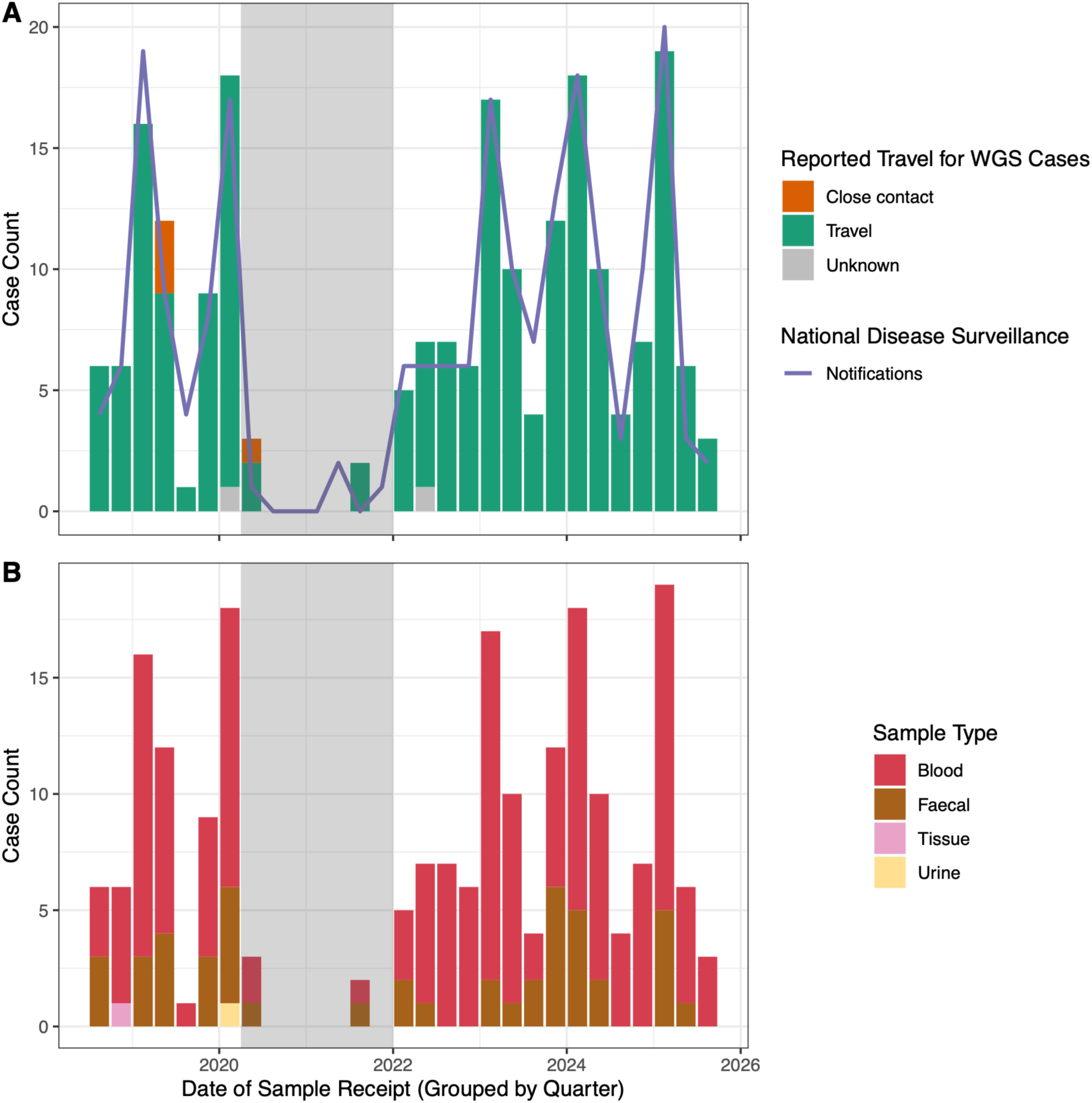
Notifications to the National Notifiable Disease Surveillance System (NNDSS) and samples received by the public health lab over time. Data is aggregated into 3-monthly year quarters. Bars represent date of sample receipt at central public health lab as the date of sample collection is not available for all samples. A) Bars are coloured by whether the patient reported recent international travel (green) or exposure to an infected close contact (orange). Patients where the infection risk could not be identified are in grey. The purple line indicates case notification rate to the National Notifiable Disease Surveillance System (NNDSS). Positive samples are identified by de-centralised hospital microbiology labs which report notifications to the NNDSS immediately while samples are received by the central public health lab at a later date. B) Bars are coloured by sample type with blood samples in red, faecal samples in brown, tissue samples in pink and urine samples in yellow. The grey box indicates period of Australian border closures due to COVID-19.

The traveller cohort was 55.1% male with a median age of 30 years (**Supplementary Figure 1**). The majority of isolates were derived from blood cultures (n=159) but were also isolated from faecal samples (n=47), urine (n=1) and tissue (n=1) (**Figure 1B**). Of the 208 unique patient cases recent overseas travel was recorded for 202 cases (**Figure 1A**). There were four cases where infection was suspected to have arisen from close contact with an infected patient (household contacts), and there were 2 cases where the source of infection could not be determined. The most frequently reported region of travel was South Asia (n=179) followed by South-East Asia (n=21) and East Asia (n=1). From returned travellers to South Asia, India was the most frequently visited country (n=150) followed by Pakistan (n=28) and Nepal (n=1) (**Supplementary Figure 2**). From traveller’s returning from South-East Asia the most frequently reported countries were Cambodia (n=11) and Indonesia (n=10). There was a single case from Hong Kong (SAR China) in East Asia. Lastly, there was a single case where recent travel was the suspected source of infection, but no region or country was recorded.

The total number of returning travellers and the country which they visited is collected by the Australian Bureau of Statistics (ABS). Between July 2018 and September 2025 (the study period) there were a total of 56.4 million travellers returning to Australia with 15.6 million returning to the state of Victoria. Regions and countries of travel are not provided at the state level and are only reported as an aggregate for all states and territories. For all Australia, South-East Asia accounted for the greatest proportion of travellers at 29.9%, followed by Oceania & Antarctica at 18.5% and North-East Asia at 12.7%. The countries most frequently travelled to were Indonesia, New Zealand and the United States of America at 13.1%, 12.7% and 7.6% respectively. We assumed that travel preferences for Victorians would mirror that of the whole nation and normalised our case count to the estimated number of Victorian travellers to each country. While India was the most frequently reported country in our dataset, Pakistan had a higher incidence per traveller with 36.0 cases per 100,000 returned travellers compared to India’s rate of 20.8 cases per 100,000 travellers. Cambodia had a similar rate to India with 19.5 cases per 100,000 travellers. Indonesia and Nepal had lower rates with 0.5 and 1.6 cases per 100,000 travellers respectively.

### Population structure of SPA

To investigate the population structure of the SPA isolates collected from returned travellers we constructed a maximum likelihood core genome phylogeny (**Figure 2**). Recombination and phage regions were masked in the core genome alignment, resulting in an alignment of 4.5Mbp with 2,101 variant sites. This analysis was supplemented with MLST using the Achtman scheme, as well as the Paratype genotyping scheme. From the 208 isolates, we identified five distinct STs with the most common being ST85 (n=95) followed by ST129 (n=91). The remaining isolates were ST1938 (n=13), ST1939 (n=5) and ST7555 (n=4). ST85, ST1938, ST1939 and ST7555 are all singe locus variants of ST129.

**Figure 2.**
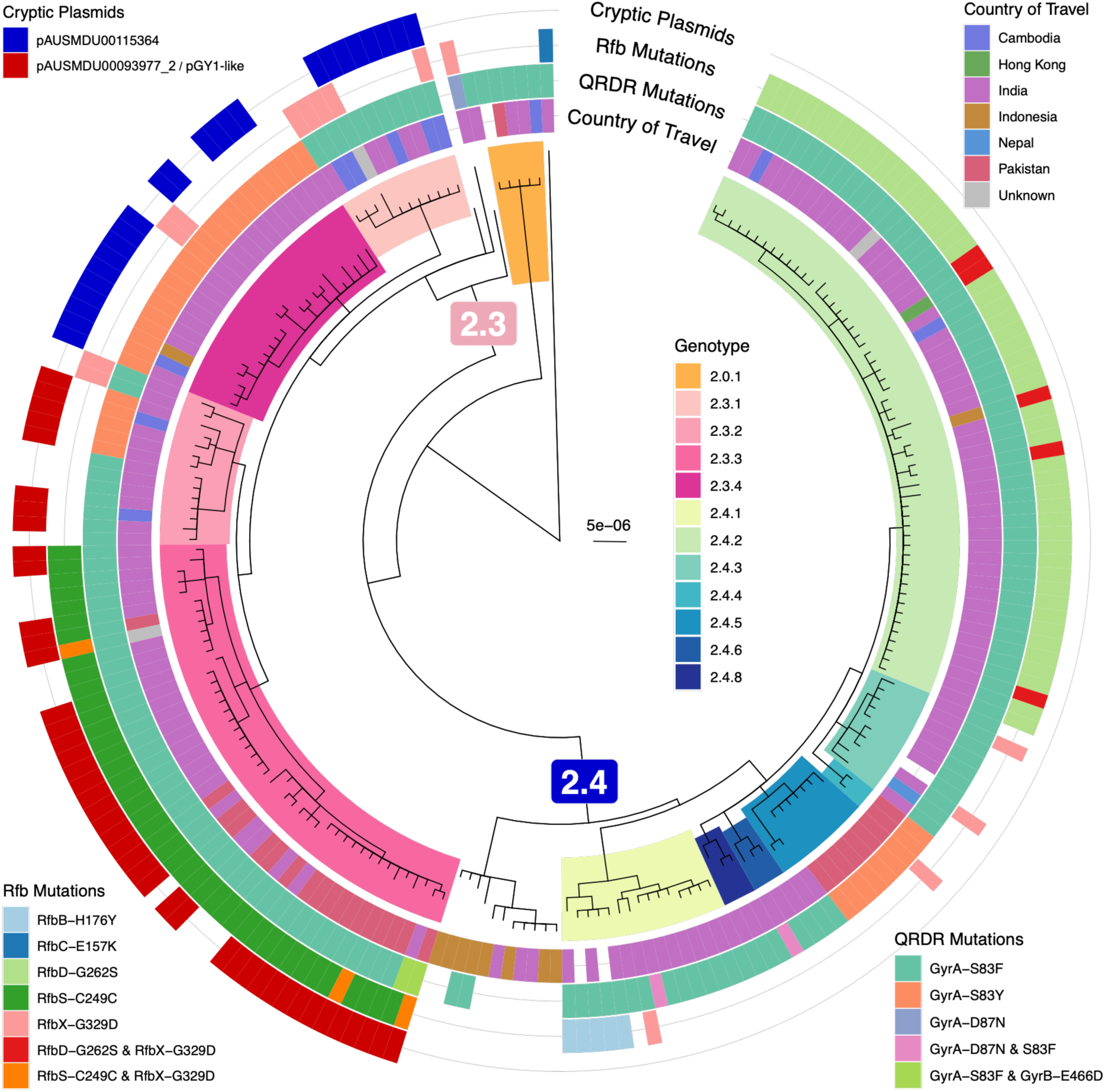
Maximum likelihood phylogenetic tree based of a core genome alignment against the SPA AKU_12601 reference genome. Clades with a Paratype genotype are coloured with genotype 2.3 in shades of pink and genotype 2.4 in shades of blue. Genotype 2.0.1 is in orange. The branches separating subclades 2.3 and 2.4 are annotated with a pink and blue box respectively. The tree is annotated with metadata in concentric rings, starting from the innermost ring is country of travel, followed by QRDR mutations, then Rfb mutations and finally presence of cryptic plasmids on the outermost ring.

A total of fifteen unique genotypes were identified in the dataset (**Supplementary Figure 3**). There was a single isolate belonging to Clade 1 (genotype 1.2.2) in a traveller returning from India. All other isolates belonged to Major Clade 2 with the most frequently reported country of travel being India. The most frequent genotypes were 2.4.2 (total=55, travel to India=50), 2.3.3 (total=45, India=26), 2.3.4 (total=22, India=20), 2.4.1 (total=15, India=13) and 2.3.2 (total=14, India=12) (**Supplementary Figure 3**). Genotype 2.4.4 was the only genotype that was not observed in a traveller returning from India, instead being found in a returned traveller from Pakistan. Genotype 2.3.3 had a notably high number of patients reporting travel to Pakistan (40%, 18/45). Both genotype 2.3.3 and genotype 2.4.1 were only observed in traveller’s returning from South Asia, whilst the other common genotypes (2.3.4, 2.3.2 & 2.4.2) were found in travellers from South and South-East Asia. We observed eleven isolates which were assigned to genotype 2.4 with no subclade given, these isolates were predominantly from travellers returning from South-East Asia, specifically Indonesia (n=8), but were also observed in traveller’s returning from India (n=3). These isolates form a distinct clade in the tree indicating support for a separate genotype designation (**Figure 2**).

### Antimicrobial resistance gene prevalence

We investigated the presence of both acquired AMR genes and point mutations with abriTAMR (41). In our unique patient dataset of 208 samples, we observed no incidence of acquired AMR genes. There were nine isolates that also lacked any resistance point mutations, suggesting they were pan-susceptible. All nine were from the unassigned subclade within genotype 2.4 that was associated with reported travel to Indonesia (**Figure 2**, **Figure 3A and Supplementary Figure 4**). Point mutations were present in the remaining 199 isolates in the remaining fourteen genotypes, with point mutations in GyrA detected in 95.7% (n=199/208) (**Figure 3A**). The most frequent was the GyrA-S83F mutation in 77.9% (n=162/208) isolates, followed by the GyrA-S83Y mutation which was present in 17.3% (n=36/208) isolates. Both GyrA-S83F and S83Y were observed in isolates from travellers returning from India, Pakistan and Cambodia, while the GyrA-S83F mutation was absent from China and Nepal (**Figure 3**). The GyrA-S83Y mutation showed strong association with genotypes 2.4.5 and 2.3.4 being present in 77.8% (n=7/9) and 100% (n=22/22) of isolates respectively. The GyrA-D87N mutation was detected in only 3 isolates which came from 3 distinct genotypes but all in cases where travel to India was reported (**Supplementary Figure 4B**). The GyrA-D87N mutation co-occurred with the S83F mutation in two of these isolates and on its own in a single isolate (AUSMDU00091465). Mutations in GyrB were rare being detected in only two of the 208 isolates. Both had the GyrB-E466D mutation as well as the GyrA-S83F mutation, both isolates belonged to genotype 2.3.3 with recent travel to India and Pakistan. No mutations in ParC/ParE were detected, nor did we observe any mutations in AcrB, which have been associated with azithromycin resistance in *S.* Typhi (62) and SPA (63).

**Figure 3.**
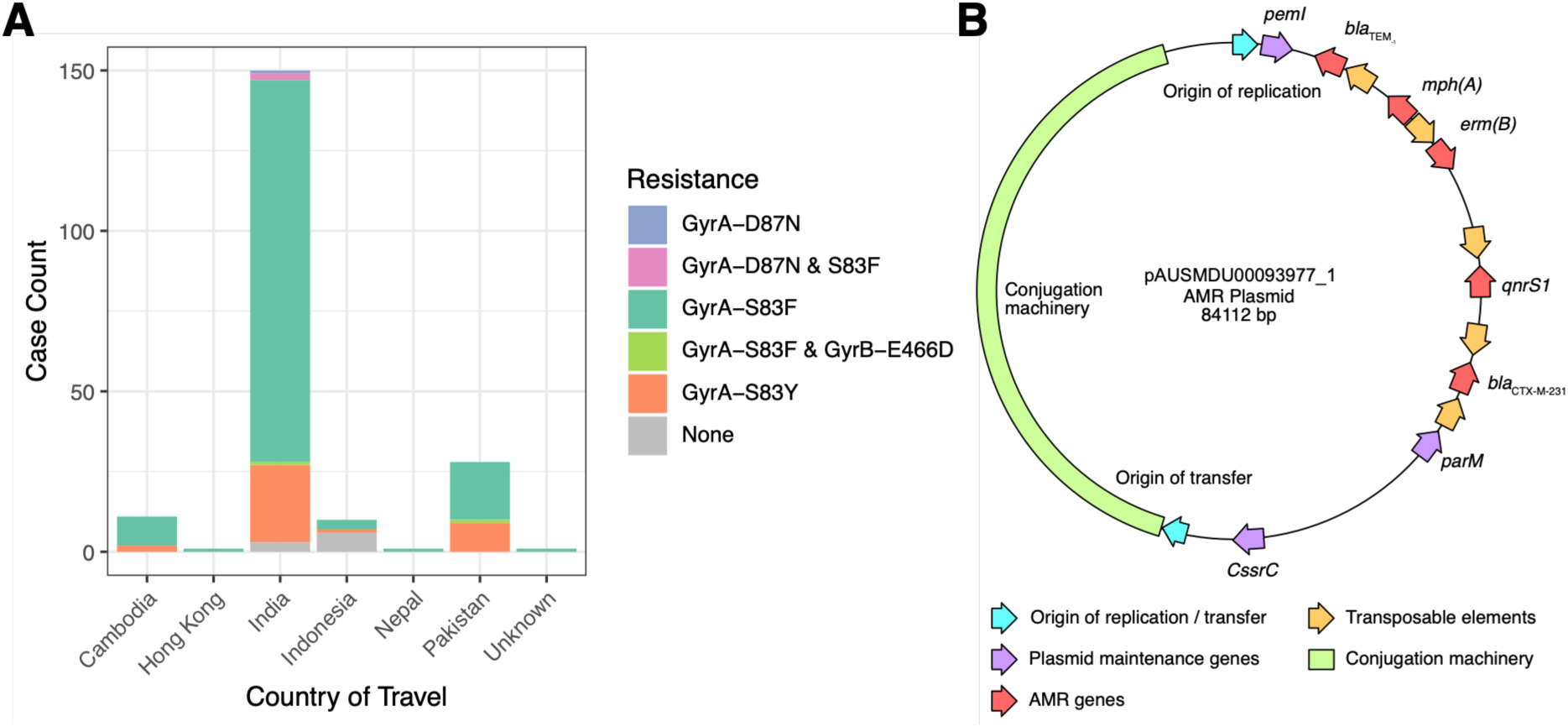
Incidence of DNA gyrase mutations and AMR plasmid map. A) Bar plot of resistance mutations in GyrA/B, by country of reported travel. B) Illustrative plasmid map of the AMR plasmid (pAUSMDU00093977_1) carried by a single isolate, not to scale. Only select genes are shown. Origins of transfer and replication in light blue, plasmid maintenance systems (partitioning and toxin / antitoxin genes) in purple, AMR genes in red, transposable elements (insertion sequences and transposons) in orange and the conjugation genes (*tra* and *trb* genes) in green.

### SPA carrying plasmid encoding an ESBL and Azithromycin resistance gene

In a single patient sample from 2023 referred to MDU PHL, a repeat SPA isolate was genomically characterised from the second colony pick from the original sample, as it had a discordant antimicrobial susceptibility testing profile compared to the first colony pick. The patient concerned had reported recent travel to Pakistan and both isolates were collected on the same day from the same faecal sample. The first isolate, AUSMDU00093461, included in the analysis above, was susceptible to amoxicillin-clavulanic acid (MIC: 2mg/L), ampicillin (MIC: 2mg/L), azithromycin (MIC: 4mg/L) and ceftriaxone (MIC: <=0.25mg/L) but resistant to ciprofloxacin (MIC: 0.5mg/L). The second isolate, AUSMDU00093977, from the second colony pick, phenotyped as extensively drug-resistant (XDR), it was resistant to amoxicillin-clavulanic acid (MIC: 16mg/L), ampicillin (MIC: >32mg/L), azithromycin (MIC: >64mg/L), ceftriaxone (MIC: 64mg/L) and ciprofloxacin (MIC: >4mg/L). As the resistant isolate, AUSMDU00093977, was the second colony pick, it was initially excluded from the genomic epidemiological investigation. However, due to its novel XDR phenotype, the AMR profile was explored in more detail. Genomic analyses revealed that while the first isolate, AUSMDU00093461, only possessed the GyrA-S83F point mutation, the second isolate, AUSMDU00093977, carried the quinolone resistance gene *qnrS* in addition to the GyrA-S83F point mutation, a beta-lactamase *bla*_TEM-1,_ an ESBL gene *bla*_CTX-M-231_, the azithromycin resistance gene *mphA*, and a macrolide resistance gene *ermB*. Both isolates were assigned genotype 2.3.3, and a core genome alignment between the two isolates confirmed that there was only a single SNP difference between the two genomes.

Long-read sequencing identified two plasmids present in the XDR isolate, AUSMDU00093977, that were absent in the non-XDR isolate AUSMDU00093461. The first was an 84.2kbp plasmid (pAUSMDU00093977_1) and the second was a 3.59kbp plasmid (pAUSMDU00093977_2). AMR gene detection with abriTAMR confirmed that the 84.2kbp plasmid (pAUSMDU00093977_1) encoded *bla*_TEM-1_, *mph(A)*, *erm(B)*, *qnrS1* and *bla*_CTX-M-231,_ while the smaller plasmid encoded no AMR or virulence genes. Gene annotation of pAUSMDU00093977_1 identified conjugative elements (*tra* / *trb* operon), toxin-antitoxin systems (*pemIK* and *sok*) and multiple transposon / insertion sequences (**Figure 3B**).

Searching the NCBI non-redundant database with BLASTn for matches to the plasmids identified an exact match (100% identity, 100% query coverage) between the 3.59kbp plasmid (pAUSMDU00093977_2) and plasmid pGY1 (EF150947.1, 3.59kbp) reported from an SPA isolate collected in 2005 (64). The best match to plasmid pAUSMDU00093977_1 (84.2kbp AMR plasmid) was pCPNKP02_AA190 (CP193630.1, 99.87% identity, 96% query coverage, 88.7kbp) found in a *Klebsiella pneumoniae* isolate collected in 2025 in Ontario, Canada (**Supplementary Figure 5**). There were five additional matches to the AMR plasmid, all above 99.8% identity and 90% coverage from a range of pathogens including *Escherichia coli* (CP090572.1), *Shigella flexneri* (CP128222.1), *Klebsiella variicola* (CP181795.1), *Shigella sonnei* (CP099773.1) and another *K. pneumoniae* strain (CP127515.1). The corresponding samples had been collected in China, Canada, Germany, Belgium and the United States of America, with collection dates ranging from 2014 to 2021 (**Supplementary Table 2**).

### Small plasmid maintained in SPA populations

Given our discovery of an MDR plasmid (pAUSMDU00093977_1) and a stably maintained small cryptic plasmid (pGY1-like, pAUSMDU00093977_2) in a single isolate we investigated whether any other plasmids were present in our dataset. There have been previous reports of SPA carrying small cryptic plasmids (65). No plasmid replicons were identified in the 208 SPA genomes, however plasmid associated genes, such as replication (*rep*) and transfer origin (*oriT*) genes, were identified in 35.1% (n=73/208) of the isolates (**Supplementary Table 3**). Three distinct gene patterns were most common in the data; replication gene accession CP000974 and OriT gene accession CP016722 (n=43), replication gene accession KU302809 (n=13), and replication gene accession EF088686 (n=11). The remaining six isolates were split across four accession gene profiles and were not investigated further.

A single representative isolate from each of the three plasmid profiles underwent ONT sequencing, assembly and BLASTn comparison with the publicly available plasmids in the NCBI non-redundant nucleotide database. The plasmid from the first accession gene profile (CP000974 / CP016722) was identical to plasmid pAUSMDU00093977_2 / pGY1-like previously identified. While the second and third accession gene profiles (KU302809 and EF088686) were in fact identical plasmids which we will refer to here as pAUSMDU00115364. These two small plasmids had matches to publicly available plasmids from other species in the Enterobacterales including *K. pneumoniae, Salmonella enterica* serovar Newport and *Escherichia coli* (**Supplementary Table 4**). Using the contigs from the long-read assemblies as query sequences we searched all 208 genome assemblies with BLASTn, restricting to matches with over 90% query coverage and 90% sequence identity.

We recovered pAUSMDU00093977_2 (pGY1-like) plasmid in 43 genomes from genotypes 2.3.2 (57.1%, n=8/14) and 2.3.3 (77.8%, n=35/45), with reported recent travel to India (n=25), Pakistan (n=15) and Cambodia (n=2). The source of infection was unidentified for the remaining patient (**Figure 2**). Annotation of pAUSMDU00093977_2 (pGY1-like) identified three coding sequences (CDS): the antitoxin *mazE*, a replication initiation protein and an unknown Nuclease-Related Domain (NERD) containing protein, in addition to an origin of transfer and a non-coding RNA (RNAI). The small pAUSMDU00115364 plasmid was detected in the draft genome assemblies of 24 isolates from genotypes 2.3.1 (72.7%, n=8/11) and 2.3.4 (72.7%, n=16/22), with patients reporting travel to India (n=18), Cambodia (n=4) and Indonesia (n=1), with one patient that had no country recorded (**Figure 2**). Annotation of pAUSMDU00115364 also identified 3 CDS, two were hypothetical proteins and the remaining was a transcriptional regulator *marR*, in addition to a non-coding RNA (RNAI).

### Diversity in point mutations and gene copy number in *rfb* locus

In our dataset we observed mutations in *rfbB, C, D, S* and *X* genes associated with the biosynthesis of the O-antigen loci. Point mutations in in this locus were strongly associated with genotype (**Figure 2**). Several mutations were only detected in a single genotype. These included the RfbB-H176Y in genotype 2.4.1 (33.3%, n=5/15), the RfbC-E157K mutation the single genotype 1.2.2 genome, the RfbD-G262S mutation in 2.4.2 (100%, n=55), and the RfbS-C249C mutation that was restricted to 2.3.3 (22.2%, n=10/45). In contrast, other mutations such as RfbX-G329D (n=22/208) were detected in different genotypes, indicative that they had emerged in multiple genetic backgrounds.

As previous studies reported that duplication of genes in the *rfb* locus were associated with changes in surface antigen structure and not point mutations in the *rfb* locus (66), we undertook ONT sequencing of representative isolates from the common genotypes in our collection to explore the differences in the O-antigen region. The *rfb* locus was completely resolved for seven isolates representing genotypes 2.0.1, 2.3, 2.3.1, 2.3.3, 2.4, and two isolates from 2.4.2. In the SPA AKU_12601 reference genome (GCF_000026565.1) there is 1,578bp between the end of *rfbX* and the start of *rfbU*, this region is annotated with a single glycosyltransferase (RefSeq: WP_011233008.1). In *S*. Typhi the gene at this position is annotated as *rfbV* however the SPA version of the gene shares only 54.08% amino acid identity with the *S*. Typhi sequence (NP_461032.1). The *rfb* locus from the seven genomes from this study were aligned to the *rfb* locus from the SPA reference genome (AKU_12601), which revealed varying levels of tandem sequence duplications between *rfbX* and *rfbU* (**Figure 4**). In genotypes 2.0.1, 2.3 and 2.4 the sequence between *rfbX* and *rfbU* had expanded to 4,463bp, while in genotypes 2.3.1 and 2.3.3 the distance had expanded to 10,245bp. The expansion was caused by a tandem duplication of a 2,885bp region from nucleotide 712 of *rfbX* to position 720 of *rfbU* (GCF_000026565.1, nucleotides 870332 to 873216). This duplicated region contained a full copy of the SPA *rfbV* gene as well as fragments of *rfbX* and *rfbU*. The tandem nature of the duplication resulted in gene fusions of *rfbU* and *rfbX* (RfbU::X) (**Figure 4**). We observed heterogeneity in genotype 2.4.2, with a single duplication event in one isolate (AUSMDU00030826) and two duplications in another (AUSMDU00025114). Genotypes 2.3.1 and 2.3.3 possessed four copies of *rfbV* and its surrounding sequence (**Figure 4**). The tandem duplications produced mixed gene annotation results from Bakta, with a variety of RfbU::X fusion products reported as glycosyltransferases (various accessions) alongside RfbU fragments.

**Figure 4.**
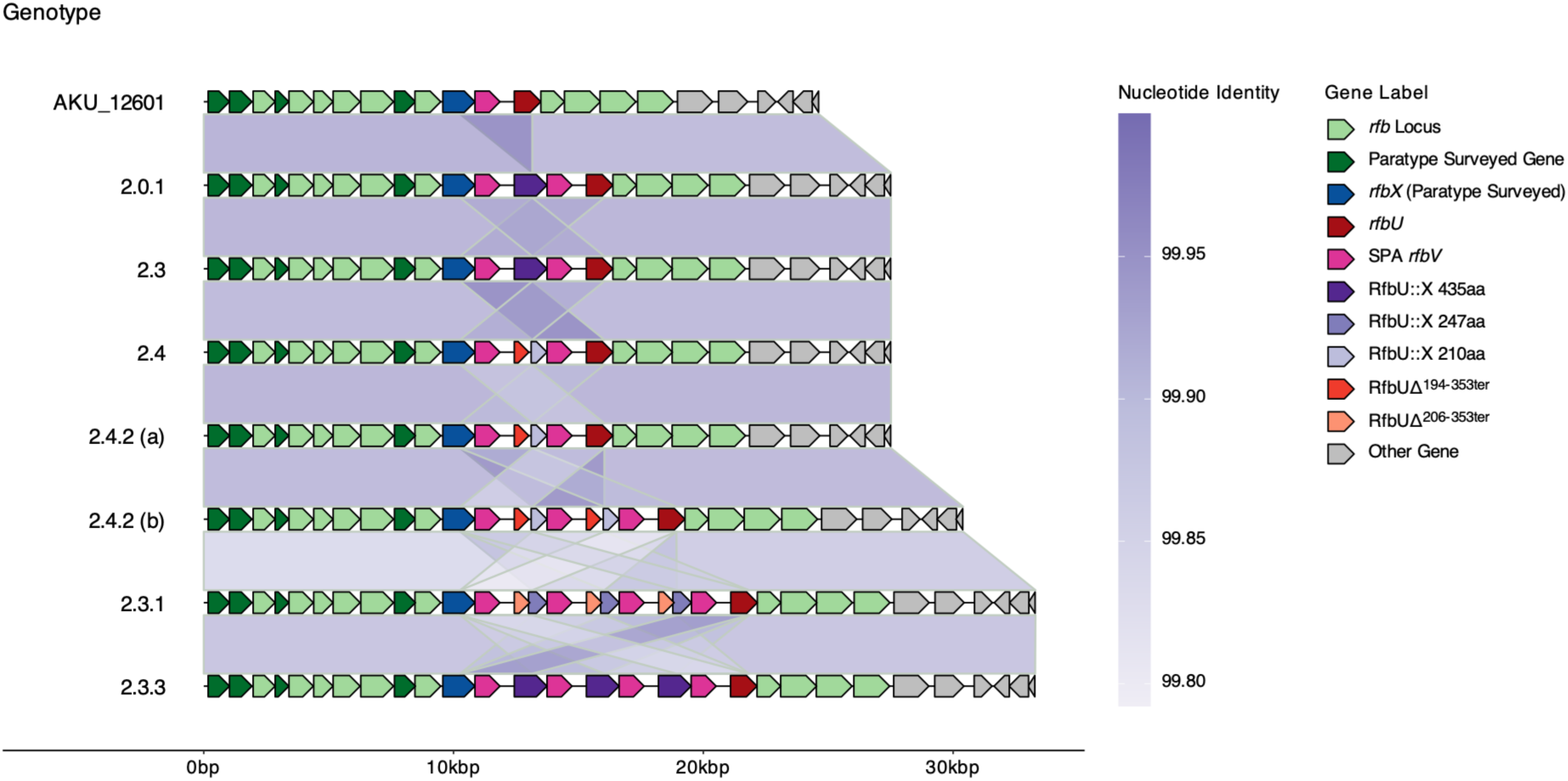
Variation in the *rfb* locus. *Rfb* locus alignment between the SPA reference genome (AKU_12601) and our closed genomes created as part of this study. Genes are annotated with arrows. Genes part of the *rfb* operon are in green, genes where Paratype reports SNPs are coloured in dark green. Paratype also reports SNPs in *rfbX* (shown in dark blue). RfbU is shown in brown and the SPA *rfbV* is shown in dark pink. RfbU and RfbX protein fusions (RfbU::X) are shown in purple, 3 different gene fusions were annotated by Bakta: a 435 amino acid gene, a 247 amino acid gene and a 210 amino acid gene. Bakta also identified RfbU protein fragments (RfbUΔ, shown in shades of orange) where the terminal portion of the protein was deleted. Two RfbU truncations were observed, a deletion from amino acid 194 to the end, and a deletion from 206 to the end. Neighbouring genes that are not part of the *rfb* locus are in grey. Sequence identity between the loci is shown with connecting purple boxes, shaded by percentage nucleotide identity.

We investigated if the differences observed in the ONT closed genome could be extended across the full dataset with short read data using an approach from a previous study that used sequencing depth to infer gene copy number. We calculated sequencing depth across the *rfb* locus and normalised to the average depth across the entire genome, however our results were inconclusive (**Supplementary Material, Supplementary Figures 6-7**).

## DISCUSSION

Here we leverage that SPA is not endemic in Australia to perform informal sentinel surveillance on a range of geographic regions frequented by Australian travellers, as has been used for *S.* Typhi (30). Our genomic dataset was able to capture all the notified cases in the state of Victoria from mid-2018, when routine sequencing of all referred *Salmonella* started, to late 2025. In total we investigated 208 unique patient cases and the travel data collected by the Department of Health and Australian Bureau of Statistics. We observed that travel to South and South-East Asia was high-risk for Paratyphoid fever, especially travel to Pakistan. This is consistent with the key epidemiological risk factors for Australian cases of typhoid fever (2,4). We had no cases from Sub-Saharan Africa, another high endemicity region however this was likely due to low rates of travel from Australia to this region.

Our observed genotypes largely align with previously reported regional geographic associations (27,67–70). Genotypes 2.4.2 and 2.3.3 were the most frequently observed in our dataset, consistent with their prior identification as globally disseminated lineages (70). There was some variation in specific country-genotype associations. In our analysis genotype 2.3.1 was acquired from travellers to Cambodia and India, however previous reports have identified this genotype predominantly in Cambodia (27,67,69), indicating continued regional spread of this genotype. Similarly, there is some discrepancies for genotype 2.4.1 which we observe mostly in travellers to India, while others have reported it mostly in samples from Nepal (69), and others have observed it across both countries (67).

Indonesia had a high rate of travel from Australia reflecting travel preferences of Australians. While Indonesia represented a low-risk region, we did observe an undefined genotype within subclade 2.4 in these isolates, consistent with previous reports of travellers returning from Indonesia (69). These isolates form a distinct clade in the phylogenetic tree, branching early within the 2.4 genotype, again consistent with previous analyses (69). This defined clade was absent from the original Paratype analysis behind the genotyping scheme, likely caused by underrepresentation of Indonesian isolates. Concerningly previous surveillance has observed this clade to establish itself within Taiwan causing local transmission (69), and this clade may warrant a defined genotyping SNP for future monitoring.

Overall, the prevalence of known AMR mechanisms in the SPA population was low, with the majority of isolates only displaying point mutations in QRDRs, consistent with other genomic surveillance studies (68,71,72). Our undefined Indonesian-associated 2.4 clade represented the lowest incidence of AMR, with many of the isolates lacking any resistance mechanisms, again consistent with previous observations (69). No AMR mechanisms were detected to ampicillin, chloramphenicol or sulfamethoxazole-trimethoprim, which have been previously, although rarely, reported by others (68,71,72), suggesting that SPA remains susceptible to older generation antibiotics. Moreover, the development and successful deployment of vaccines targeting *S*. Typhi is anticipated to reduce the need for antibiotic therapy, further supporting a possible return to older generation antibiotics.

Notably, we did observe a single isolate carrying a plasmid encoding both ESBL and azithromycin resistance genes. While occurrence of ESBL genes in SPA isolates has been reported before (73,74), to our knowledge this is the first report of gene mediated azithromycin resistance in SPA. However, the *mphA* resistance gene has been observed in non-Typhoidal serovars and is often mobilised on plasmids (75). The occurrence of plasmid mediated resistance in SPA is concerning, as it could follow the evolutionary path as *S*. Typhi where a resistance cassette moved from carriage on plasmids, to integrating into the chromosome and becoming fixed in several genotypes (14,76). We observed several mobilizable elements on the MDR plasmid which could facilitate this genomic transition. We could not determine whether the discordant AST results from the duplicate colony picks were due to spontaneous plasmid acquisition or plasmid loss. However, the susceptible isolate, AUSMDU00093461, was missing both the MDR plasmid and a small cryptic plasmid. We reason that AUSMDU00093461 likely lost both plasmids during culture and sequencing, rather than spontaneously acquiring both simultaneously. The MDR plasmid characterised from AUSMDU00093977 was detected from a range of geographical locations and bacterial species, suggestive that it has been circulating in Enterobacteriaceae species since 2014, and on this instance acquired in SPA. This highlights the need for continued plasmid AMR surveillance, particularly with the changing selection pressures due to the rollout of the TCV, that may drive populations expansion of SPA.

Plasmids in SPA have been reported as early as 2003 (77), yet are not frequently reported from genomic surveillance. Previous publications have reported small plasmids in genotype 2.3 (65). The small cryptic plasmids reported here, pAUSMDU00093977_2 (pGY1 like) and pAUSMDU00115364, do not possess conjugation machinery. Given the high sequence similarity of the plasmids collected over several decades, we infer that it has been stably maintained by genotype 2.3 for decades (64). It is unclear what function, if any, these plasmids may be performing. Small cryptic plasmids have been observed in *E. coli*, removal of these plasmids from their host strains resulted in minimal transcriptional changes suggesting they posed little to no fitness cost on the cell (78).

We detected differences in the O-antigen biosynthesis *rfb* locus, including both point mutations, and gene copy number variation. SNPs in the *rfb* locus were frequently associated with different genotypes, however previous studies have indicated that these SNPs do not correlate with phenotypic changes in O-antigen structure (66). We observed that gene duplication of *rfbV* is occurring alongside a variety of *rfbUX* fusion genes, however we suspect these gene fusions are non-functional. Copy number variation was observed to occur within a single genotype suggesting this is a dynamic process in SPA, spontaneous tandem duplications have been reported in *Salmonella* (79). Future work should confirm whether copy number variation is causing surface antigen variation, specifically that increases in *rfbV* expression is causative, as well as confirming that variation is occurring within genotypes. Elucidating the dynamics of the O-antigen in SPA will be important for current and future vaccine efforts.

Our informal sentinel surveillance identified that incidence of AMR mechanisms in SPA remains low, primarily mutations in DNA gyrase enzymes contributing to fluoroquinolone resistance. With a vaccine driven decrease in *S*. Typhi this may support a return to older generation antibiotics for treatment of SPA. Concerningly however, we identified a single isolate harbouring a plasmid encoding genes conferring resistance to last-line antibiotics such as third-generation cephalosporins and azithromycin. This isolate still lacked AMR mechanisms against older generation antibiotics. We confirmed the presence of small cryptic plasmids in genotypes 2.3 that have been stably maintained for decades. Our analysis indicates these are parasitic plasmids, with little biological function. Lastly, we further showed gene copy number variation occurring in surface antigen biosynthesis genes. This variation should be investigated further due to its potential impact immunogenicity profiles and vaccine efficacy. Together these data provide a comprehensive baseline for future genomic surveillance of SPA in Australia and the surrounding region. Linking the epidemiological and genomic data to explore population structure, changing AMR profiles and differences in biosynthesis genes, will ensure emerging drug-resistant SPA are identified early and can inform public health recommendations for SPA.

## Abbreviations

SPA: *Salmonella enterica* serovar Paratyphi A

## FUNDING

The Microbiological Diagnostic Unit Public Health Laboratory is funded by the Victorian Government, Australia. BPH is supported by a Leadership Fellowship from the NHMRC of Australia (GNT2041625). DJI was supported by an Emerging Leadership Fellowship from the NHMRC of Australia (GNT2041653). RRW was supported by an Australian Research Council Discovery Early Career Researcher Award (DE250100677).

## Notes

### Competing Interest Statement

The authors have declared no competing interest.

