## Supplementary Materials for "Genomic epidemiology and emerging antimicrobial resistance profiles of *Salmonella* Paratyphi A in returning travellers to Australia"

Connor *et al.*

### SUPPLEMENTARY MATERIALS

#### SUPPLEMENTARY METHODS

##### Estimating *rfb* locus duplication from sequencing reads

Read alignment files produced by Snippy for the phylogenetic analysis (see main manuscript methods) were used to quantify sequencing depth across the genome for each isolate. Sequencing depth was quantified with Samtools depth v1.20 (1), using the BAM files and default parameters. Per base sequencing depth was aggregated, normalised and used for plotting in R. Sequencing depth was normalised to either the average (mean) depth across the full genome, or the average (mean) depth across *rfbS* (GCF_000026565.1 nucleotides 867698 to 868538)

#### SUPPLEMENTARY RESULTS

##### Matches to the cryptic plasmids in public databases

The best publicly available match for pGY1 was to the plasmid CP162194.1 (91% query coverage, 97.33% identity) from a *K. pneumoniae* isolate collected from a food sample in Switzerland. There were additional hits to plasmid sequences from multiple isolates of *S*. Newport. The best match in the core nucleotide database for pAUSMDU00115364 was CP036184.1 (94.1% identity and 94% query coverage), matching the previously reported accession (2). This sequence corresponded to a plasmid identified in an *E. coli* isolate collected in 2017 in China. There were additional full-length hits to other *E. coli* plasmid sequences such as CP145659.1 and CP148572.1 which had 99.49 and 99.34% identity, but both were considerably longer than the query plasmid (2.61kbp) at 5.7kbp and 11.4kbp respectively, possibly representing plasmid co-integrates.

##### Measuring gene duplication with sequencing depth

As a previous publication had used sequencing depth to measure duplication at the *rfb* locus we investigated if the differences observed in the ONT closed genome could be extended across the full dataset with short read data using the same approach. We calculated sequencing depth across the *rfb* locus and normalised to the average depth across the entire genome. We observed variable depth across the full *rfb* locus (**Supplementary Figure 6**), depth at the start of the locus

(GCF_000026565.1, nucleotides 860063 to 867698) averaged 0.976 indicating similar depth to the rest of the genome. Depth dropped to an average of 0.396 at the start of the *rfbS* gene (GCF_000026565.1, nucleotides 867698 to 868538). Spikes in read depth were observed across the tandem duplication we identified in our closed genomes (**Supplementary Figure 7A**). The increases in sequencing depth occurred over the terminal section of *rfbX* (GCF_000026565.1, nucleotides 870600 to 870919), start of *rfbV* (871100 to 871500) and start of *rfbU* (872496 to 873000), corresponding to regions of the 2,885bp tandem duplication we observed in our closed genomes.

The magnitude of read depth increase varied by genotype with 2.4.3 exhibiting the highest average peaks and 2.0.1 the lowest (**Supplementary Figure 7A**). Averaging the genome-normalised sequencing depth across the full 2,885bp duplication confirmed variable sequencing depth with it ranging from 0.262 to 2.450 across all genotypes (**Supplementary Figure 7B**). Focussing on the samples which had closed genomes showed that the depth pattern did not fully match our initial observations. Genotypes 2.0.1, 2.3, and 2.4 all had two copies of the 2,885bp sequence in the closed genomes, however the calculated read depth for these samples was 0.262, 0.443 and 0.511 respectively, suggesting reduced copy number relative to the rest of the genome. The two closed genomes of genotype 2.4.2 exhibited differences in duplication number (two copies vs. three), however the normalized read depth of the same two isolates suggested no differences (two copies: 0.992 vs. three copies: 1.01). As we had observed a reduction in sequencing depth at the start of *rfbS* we reasoned that sequencing coverage over this region may be poor, we explored normalizing depth across the 2,885bp duplication to the average depth of *rfbS* rather than the full genome (**Supplementary Figure 7B**). While this increased read depth above 1 for all samples, in line with expected results for a duplication event, the results still did not match the closed genomes. Specifically, genotypes 2.0.1 and 2.4 had depths closer to 1 rather than 2 (1.24 and 1.09), and the two isolates from genotype 2.4.2 still had similar depths (2 copies: 2.05, 3 copies: 2.10).

### SUPPLEMENTARY FIGURES

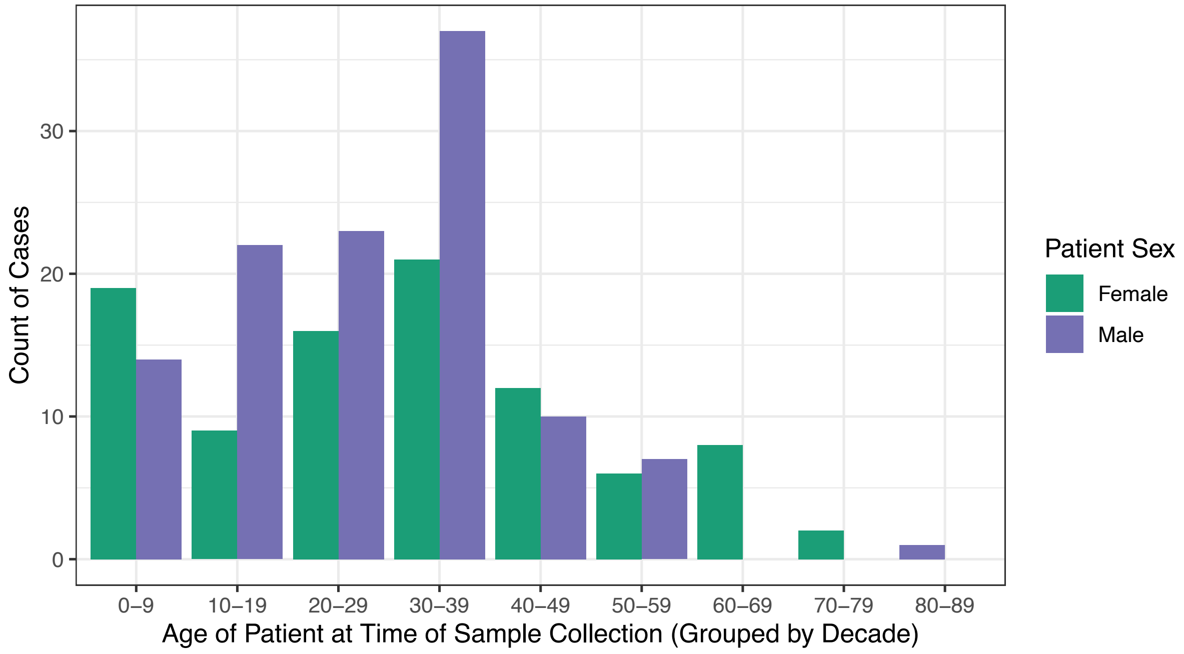


**Supplementary Figure 1. Patient age and sex at time of sampling.**

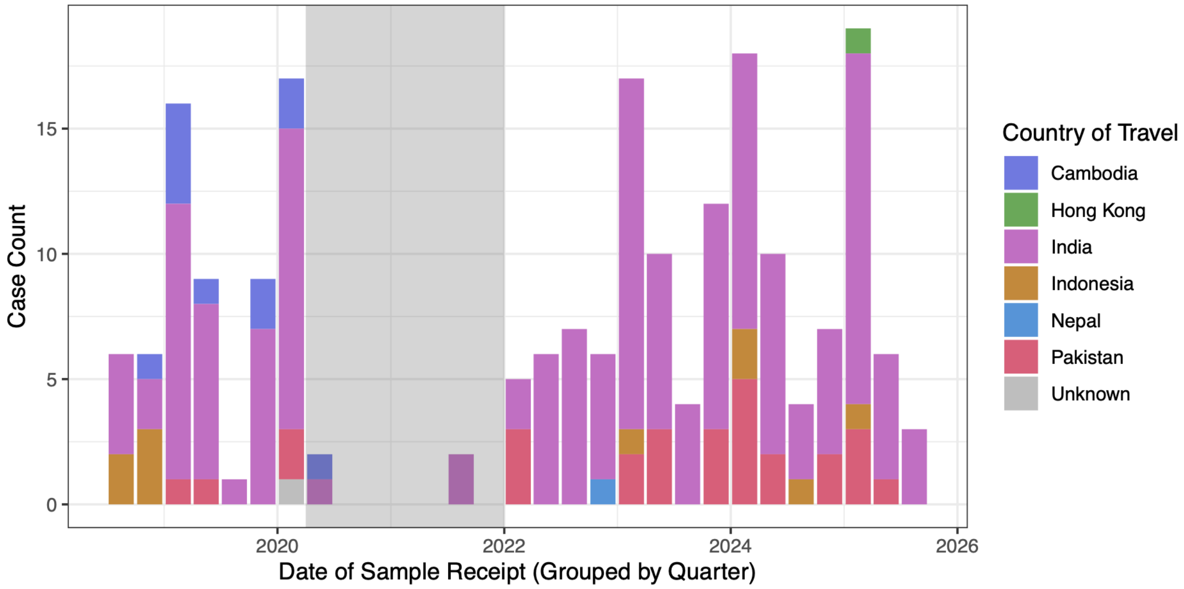


**Supplementary Figure 2. Reported country of travel over time.**
Data is aggregated into year quarters. Grey box corresponds to period of Australian border closures.

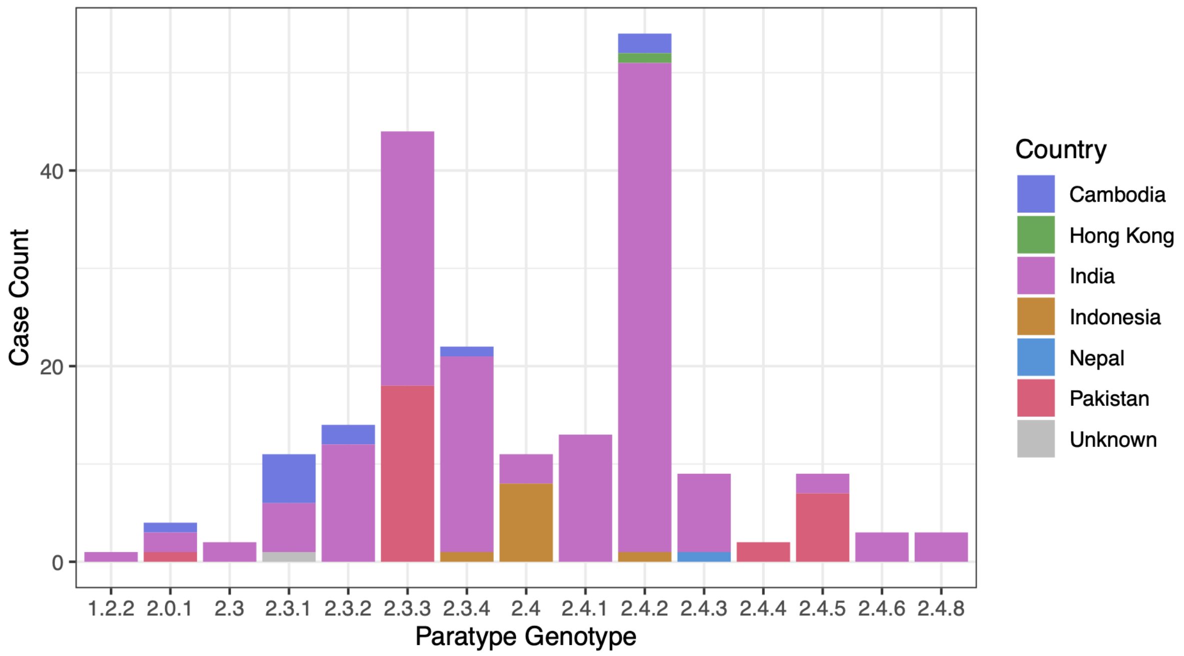


**Supplementary Figure 3. Paratype assigned genotypes coloured by reported region of travel.**

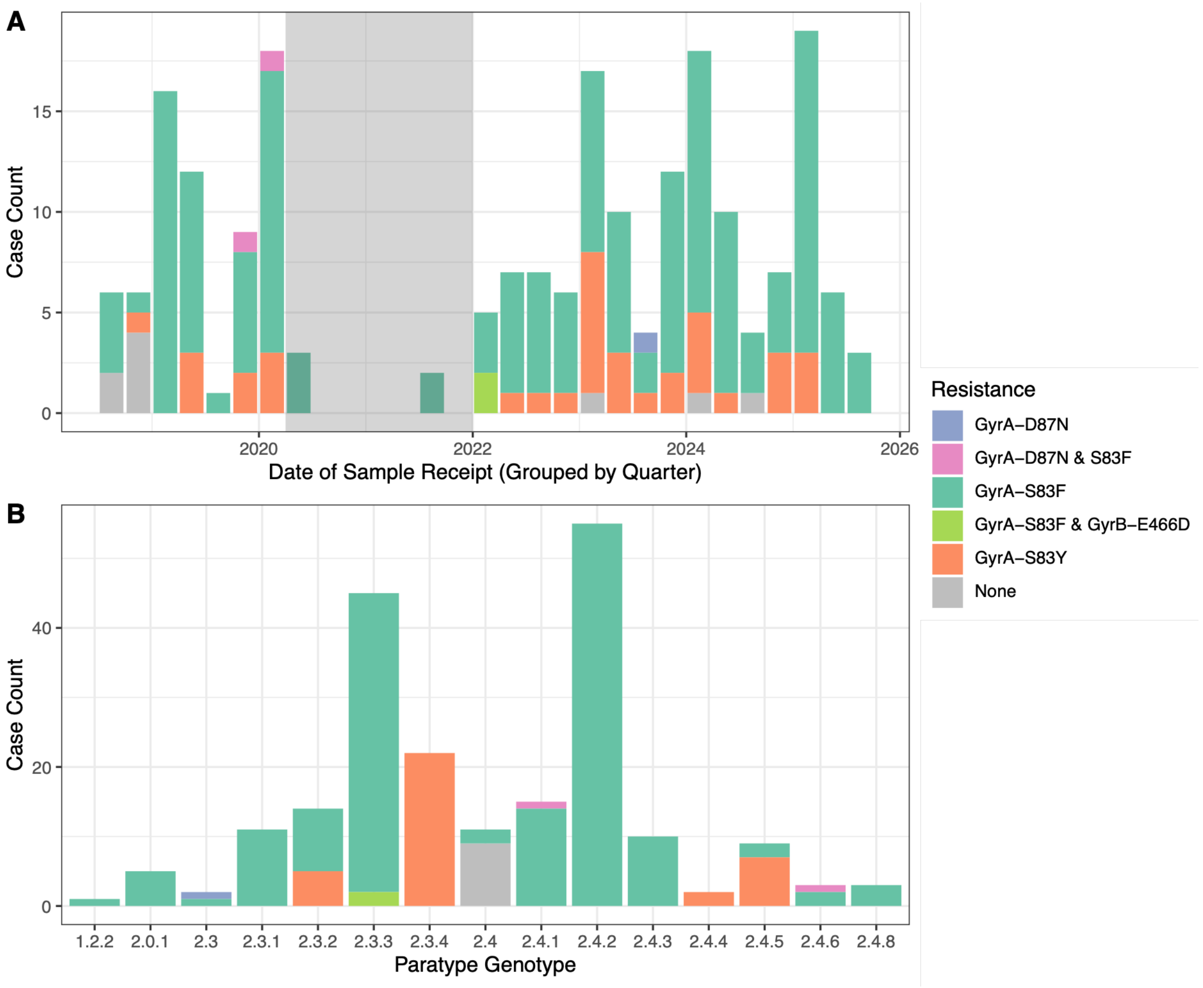


**Supplementary Figure 4. AMR mechanisms in SPA**

A) AMR mechanisms over time. Data is aggregated into quarters and bars are coloured based on AMR. B) Occurrence of AMR in Paratype assigned genotypes.

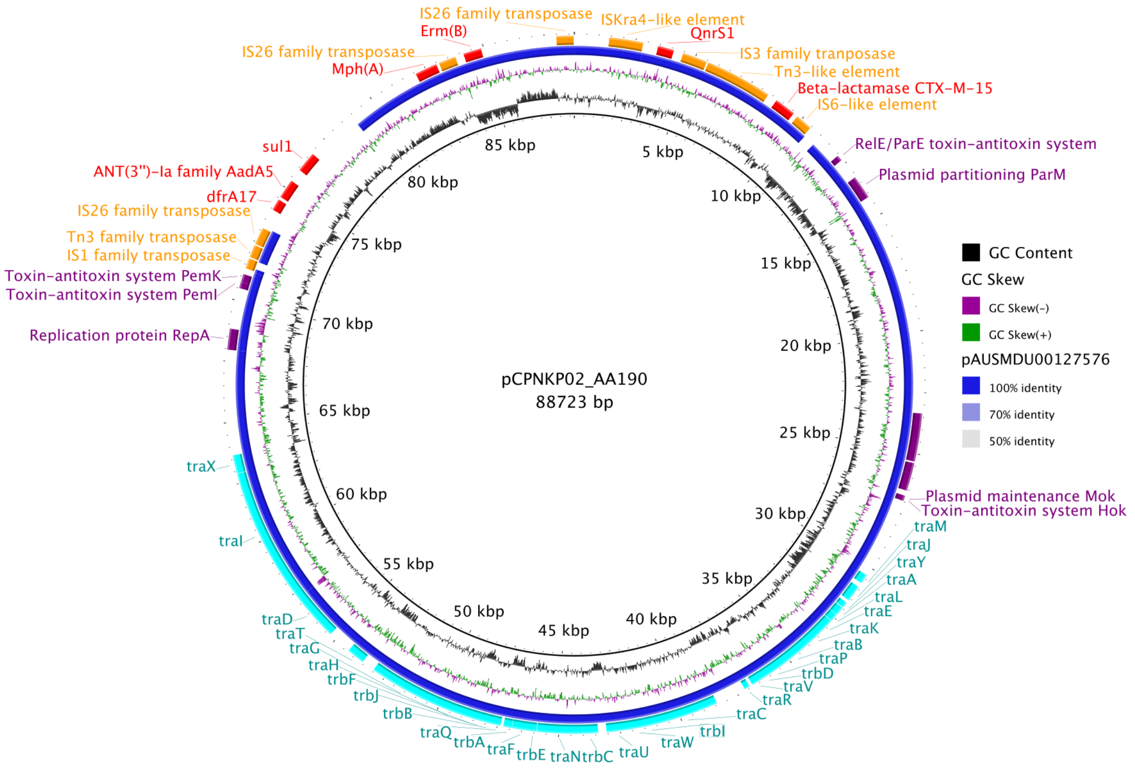


**Supplementary Figure 5. Plasmid alignment between the MDR plasmid pAUSMDU00093977 and its closed match in public databases (pCPNKP02_AA190).** The reference sequence pCPNKP02_AA190 (CP193630.1) from a *K. pneumoniae* aligned against the sequence of pAUSMDU00093977_1 (dark blue outer ring). Gaps in the outer dark blue ring indicate regions that are absent in pAUSMDU00093977_1. The GC content and GC skew of pCPNKP02 are shown in the inner rings as black and green / purple rings. Select gene annotations for pCPNKP02 are on the outer ring. Plasmid conjugations systems are in light blue, mobile genetic elements are in orange, plasmid maintenance systems in purple and AMR genes in red. Plasmid pAUSMDU00093977_1 has lost AMR genes *sul1, dfrA17* and *aadA5* (shown as gap in blue ring), but gained *bla*$_{TEM-1}$ (not shown on figure, not present on pCPNKP02). The ESBL gene is annotated as *bla*$_{CTX-M-15}$ here rather than *bla*$_{CTX-M-231}$ due to differences in annotation tools (Bakta vs. abriTAMR). The alleles for *bla*$_{CTX-M-15}$ and *bla*$_{CTX-M-231}$ differ by a single SNP.

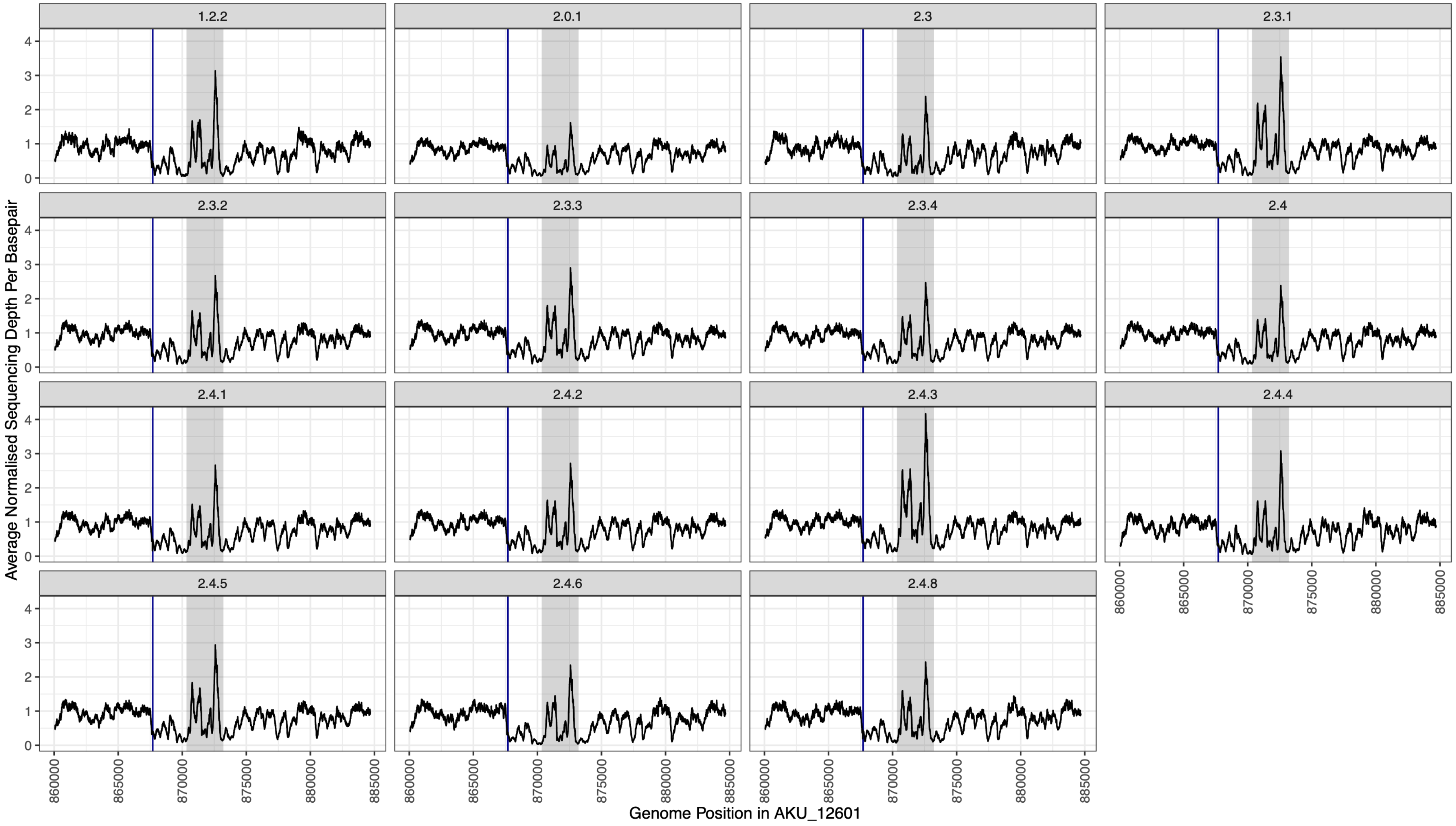


**Supplementary Figure 6. Sequencing depth, normalised to genome average depth, across the *rfb* locus for each identified genotype.** The blue line indicates the start of the *rfbS* gene, where depth drops. The grey box indicates the 2.8kbp identified tandem repeat.

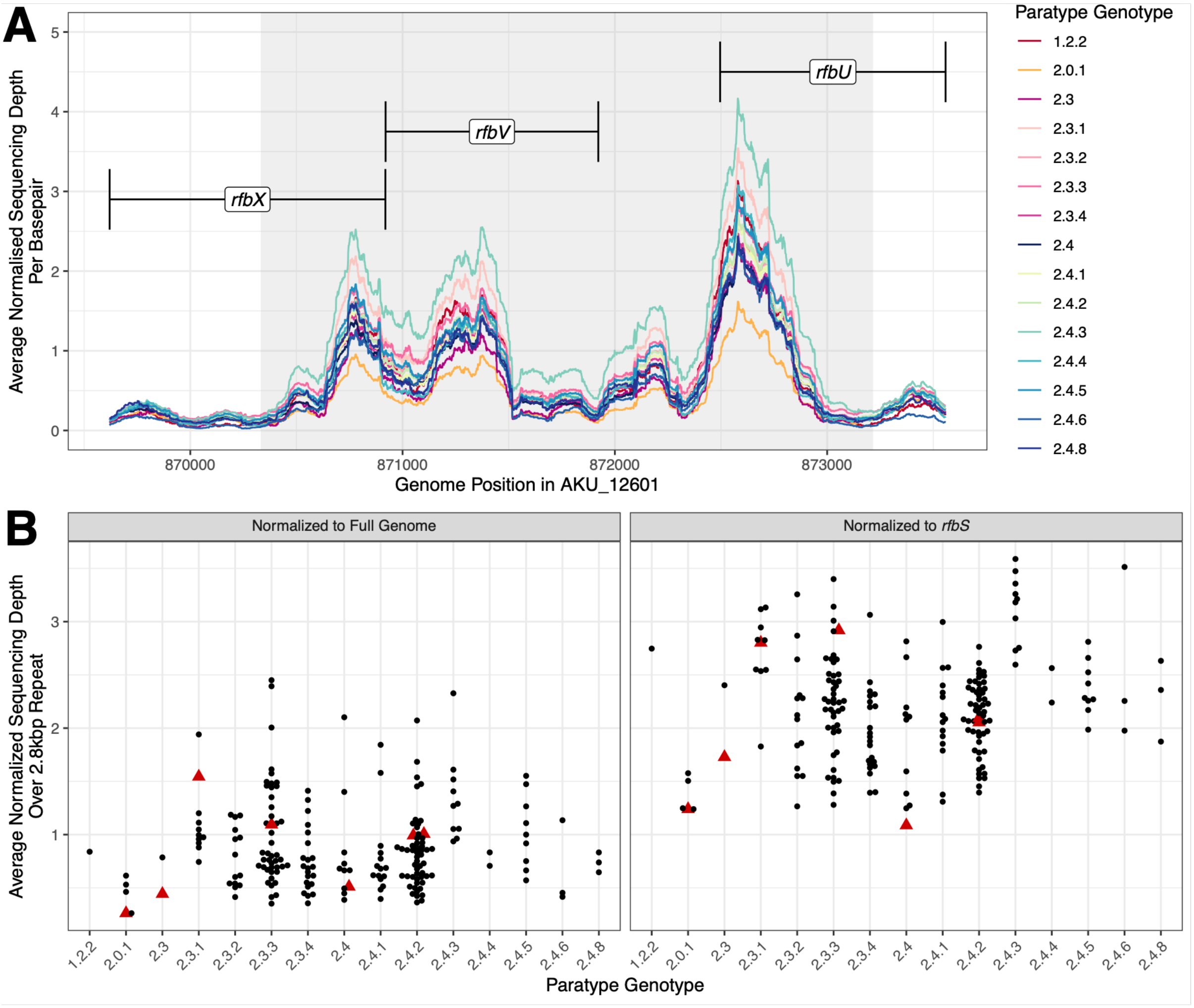


**Supplementary Figure 7. Sequencing depth across the region of duplication and aggregated sequencing depths for individual isolates.**
A) The sequencing depth per base across *rfbX*, *rfbV* and *rfbU*. Depth is normalised to the average depth across the entire genome and averaged per genotype. Lines are coloured for each genotype. B) The average sequencing depth across the 2.8kbp tandem repeat unit normalised to either the average depth across the entire genome, or to the average depth across the *rfbS* gene. Each point represents the normalised depth for an individual genome. Red triangles are the depth values calculated for the isolates for which we were able to generate closed genomes with an intact *rfb* locus.
